# Effectiveness of training general practitioners in the ABC versus 5As method of delivering brief stop-smoking advice: a pragmatic, two-arm cluster randomised controlled trial

**DOI:** 10.1101/2020.03.26.20041491

**Authors:** Sabrina Kastaun, Verena Leve, Jaqueline Hildebrandt, Christian Funke, Stephanie Klosterhalfen, Diana Lubisch, Olaf Reddemann, Hayden McRobbie, Tobias Raupach, Robert West, Stefan Wilm, Wolfgang Viechtbauer, Daniel Kotz

**Affiliations:** Institute of General Practice (ifam), Centre for Health and Society (chs), Addiction Research and Clinical Epidemiology Unit, Medical Faculty of the Heinrich-Heine-University Düsseldorf, 40225 Düsseldorf, Germany; University of New South Wales, National Drug and Alcohol Research Centre, 2031 Randwick, Australia; Lakes District Health Board, 3010 Rotorua, New Zealand; Department of Cardiology and Pneumology, University Medical Centre Göttingen, 37075 Göttingen, Germany; Behavioural Science and Health, Institute of Epidemiology and Health Care, University College London, WC1E 6BLondon, UK; Department of Psychiatry and Neuropsychology, Maastricht University, 6200 MD Maastricht, Netherlands; Department of Family Medicine, CAPHRI School for Public Health and Primary Care, Maastricht University, 6200 MD Maastricht, The Netherlands

**Keywords:** Tobacco addiction, primary care, general practitioner, brief advice, smoking cessation, clinical guideline, 5As, ABC, very brief advice

## Abstract

**OBJECTIVE:** To assess the effectiveness of a 3.5h-training for general practitioners (GPs) in two different methods (ABC, 5As) of giving brief stop-smoking advice during routine consultations.

**DESIGN:** Pragmatic two-arm cluster randomised controlled trial with pre-post-design for the primary outcome and cluster randomisation for secondary outcomes.

**SETTING:** General practices, North Rhine-Westphalia (German federal state), recruited 2017-2019.

**PARTICIPANTS:** Practices were randomised (1:1) to an ABC or 5As training. Tobacco smoking, adult patients, who consulted trained GPs in these practices in the 6 weeks prior to or following the training were eligible to participate. Ineligible were: non-smokers, patients who did not meet the GP in person, or could not provide informed consent.

**INTERVENTIONS:** Two different standardised 3.5h-trainings (ABC or 5As) for GPs in delivering brief stop-smoking advice were carried out per study cycle (six cycles in total). Trainings were moderated by a senior researcher and an experienced GP, and included role-plays with professional actors.

**MAIN OUTCOME MEASURES:** Primary outcome: patient-reported rates of GP-delivered stop-smoking advice prior and following the training, irrespective of the training method. Secondary outcomes: patient-reported receipt of recommendation/prescription of evidence-based smoking cessation treatment: including behavioural support, any pharmacotherapy (nicotine replacement therapy (NRT), varenicline, or bupropion), or a combination therapy; and the effectiveness of ABC versus 5As regarding all outcomes.

**RANDOMISATION AND MASKING:** Computer-generated block randomisation or, if not feasible, randomisation based on the GPs’ temporal availability at training dates. GPs were not fully blinded. Patients were blinded to the nature of the study until data collection ended.

**RESULTS:** 52 GP practices (27 ABC, 25 5As) with 69 GPs were included. Of 5,406 patients who provided informed consent, 1,937 (35.9%) were current smokers, of whom 1,039 were interviewed prior to and 898 following the training. GP-delivered stop-smoking advice increased from 13.1% (n=136/1,039) to 33.1% (n=297/898) following the training (adjusted odds ratio (aOR)=3.25, 95%CI=2.34 to 4.51). Recommendation/prescription rates of treatment were low (<2%) pre-training, but had increased after the training (e.g., behavioural support: aOR=7.15, 95%CI=4.02 to 12.74; any pharmacotherapy: aOR=7.99, 95%CI=4.11 to 15.52). GP-delivered stop-smoking advice increased non-significantly (p=0.08) stronger in the ABC vs. 5As group (aOR=1.71, 95%CI=0.94 to 3.12).

**CONCLUSIONS:** In GPs in Germany, a single session of training in stop-smoking advice was associated with a three-fold increase in rates of advice giving and a seven-fold increase in offer of support. The ABC method may lead to higher rates of GP-delivered stop-smoking advice during routine consultations. Approaches to further increase the delivery of such advice, and upscaling implementation strategies for the training in general practice, should be evaluated.

**TRIAL REGISTRATION:** German Clinical Trials Register: DRKS00012786.

**WHAT IS ALREADY KNOWN ON THIS TOPIC:**

- The implementation of clinical guideline recommendations stating that general practitioners (GPs) should routinely deliver brief stop-smoking advice and offer evidence-based smoking cessation treatment is low in Germany, a country with a smoking prevalence of ∼28%.
- A strategy is needed to overcome barriers (e.g., lack in knowledge and skills) preventing GPs from routinely delivering stop-smoking advice. No experimental study has evaluated such a strategy in German general practice so far.
- Two different methods of delivering such advice are recommended in the national guidelines – ABC and 5As – but it is unclear which method can be more effectively implemented by trained GPs.

**WHAT THIS STUDY ADDS:**

- This cluster randomised controlled trial evaluated the effectiveness of a 3.5h-training for GPs in delivering brief stop-smoking advice and compared two different methods (ABC vs. 5As) regarding the rates of delivery of such advice and recommendations of evidence-based cessation treatment in 1,937 smoking patients from 52 GP practices in Germany.
- The training, irrespective of the method, was associated with a three-fold increase in rates of advice giving and a seven-fold increase in the offer of support.
- The data indicate that training according to ABC may be more effective than 5As in increasing the rates of GP-delivered stop-smoking advice.

## INTRODUCTION

National and international clinical guidelines^1-4^ on the treatment of tobacco addiction strongly recommend that healthcare professionals should routinely give brief stop-smoking advice to every smoking patient and provide evidence-based smoking cessation treatment, since particularly the combination of such advice together with behavioural (e.g., outpatient individual or group intervention) or pharmacological (e.g., nicotine replacement therapy (NRT), varenicline or bupropion) therapy substantially increases long-term abstinence rates.^5 6^ Primary care physicians play a pivotal role in smoking cessation because the majority of smokers visit their general practitioner (GP) at least once a year.^7^ However, the implementation of these guideline recommendations in German general practice is rather poor. Only about 18% of smokers report the receipt of brief advice to quit smoking during their consultation, including barely 4% who report the offer of an evidence-based smoking cessation treatment.^7^ Central barriers for GPs to the provision of brief stop-smoking advice include the lack of training or education in the delivery of effective advice to quit, and the lack of time to deliver advice during routine consultations.^8-12^ Such training is not implemented by default in the medical education in Germany, and postgraduate trainings are optional and have to be paid by the physicians themselves.

Systematic review evidence supports the effectiveness of training health professionals in delivering smoking cessation advice on the point prevalence of smoking, continuous abstinence and professional performance.^13^ Accordingly, guidelines for the implementation of Article 14 of the World Health Organization’s Framework Convention on Tobacco Control (FCTC) recommend that healthcare workers should be trained to deliver such advice to their smoking patients.^14^ However, only few trials on the effectiveness of such trainings have been conducted in general practice settings.^13^ These studies show positive effects of trainings of varying durations from 40 minutes^15 16^ to several days^17^ on the rates at which GPs deliver brief stop-smoking advice,^17 18^ refer to smoking cessation services,^15^ and on GP-reported knowledge, self-efficacy, and attitude regarding the delivery of brief smoking cessation counselling.^17 19^ So far, only one cluster randomised trial on the effectiveness of a smoking cessation counselling training for GPs combined with financial incentives has been conducted in Germany^20^ but due to the design of this trial, no conclusion can be drawn regarding the unique effect of the training.

Brief stop-smoking counselling can be structured in many ways. The German clinical guideline^3^ for treating tobacco addiction recommends the provision of either the 5As method^1^ (ask for the smoking status, brief advice to quit, assess the motivation to quit, assist by providing evidence-based treatment, arrange follow-up contacts) or the much briefer ABC method^21^ (ask for the smoking status, brief advice to quit, cessation support by providing evidence-based treatment). So far, no studies comparing the effectiveness of both methods on the rates of delivery of brief stop-smoking advice in general practice settings have been published, and thus no recommendation can be made to favour one method over the other. However, the implementation of the 5As method during routing general practice seems to imply some disadvantages. According to this method, only smokers willing to quit receive the final steps “assist” and “arrange”. For those unmotivated to quit, which applies to most smokers at a consultation,^22,23^ GPs are recommended to provide an additional intervention to enhance the motivation to quit (the 5Rs^1^), which may lead to time-consuming and frustrating discussions. Several studies^24 25^ showed that, probably due to the length of this method, the last two steps of 5As are only rarely applied, although associations between these steps and success in quitting is strongest,^26^. Thus, many smokers would not be offered evidence-based treatment to quit smoking, and may be less likely to use such treatment when feeling motivated and attempting to quit at a later stage. Since it may be assumed that the ABC method is more convenient to apply for GPs in daily practice, we developed and pre-tested two 3.5h-trainings for GPs in delivering such advice during routine consultations: one based on the 5As and one based on the ABC method.^27^ The aim of the present study was to assess whether these trainings provide an effective strategy to improve the implementation of the clinical guideline recommendations for the treatment of tobacco addiction by increasing patient-reported rates of GP-delivered brief stop-smoking advice (primary outcome) and the recommendation/prescription rates for evidence-based smoking cessation treatment (secondary outcomes). A secondary aim was to compare the effectiveness of ABC and 5As against each other.

## METHODS

### Trial design

Detailed information on the development and pilot testing of the intervention, and on the results of a qualitative process evaluation have been published previously in a protocol.^27^ Non-adherence to this study protocol, if applicable, will be reported throughout this manuscript and extensions to planned analyses will be reported under the corresponding “non-adherence” paragraph. We conducted a pragmatic, two-arm cluster randomised controlled trial with a pre-post-design for the primary outcome (evaluation of the effectiveness of a training on the delivery rates of brief stop-smoking advice during routine consultations) and with cluster randomisation for the comparison of the effectiveness of both training methods – ABC and 5As – against each other. The study protocol was approved by the medical ethics committee at the Heinrich-Heine-University (HHU) Düsseldorf, Germany (5999R). All participants gave written informed consent. GP practices were randomised between 22.06.2017 and 15.03.2019. The study had been registered at the German Clinical Trials Register (DRKS00012786).

The study consisted of six cycles. Initially, according to the study protocol, a study cycle was defined as a period of eight weeks: 4 weeks pre-training data collection followed by the training (intervention) and then 4 weeks post-training data collection. In order to minimise sampling bias which might be aligned with considerable fluctuations in patient flow among different practice days or weeks (e.g., on Mondays, during a flu epidemic, or on public holidays), data collection should be carried out on about seven varying GP office days during the four-week pre-training period and on seven days during the post-training period. However, these periods had to be extended to up to 6 weeks with up to 10 days of data collection due to, e.g., lower daily patient visits, practice holidays, or difficulties to schedule dates for data collection. Per cycle, two trainings were carried out, one according to each method.

### Participants

#### GP practices

GP practices were recruited by postal dispatch from the publicly accessible online medical register of the regional Association of Statutory Health Insurance Physicians North Rhine in the Rhine-Ruhr Metropolitan Region of the German federal state North Rhine-Westphalia: a densely populated, polycentric urban agglomeration area with a strongly intercultural shaped population structure and economic inequalities, as well as from the practice network of the Institute of General Practice of the HHU Düsseldorf, Germany. Practices interested in participation were contacted by phone and fax messages in all following conversations. According to the study protocol,^27^ all GPs from group or single practices were eligible except for those specialised in treating substance abuse or in psychotherapeutic care, or those who have been trained in delivering smoking cessation support within the last five years. However, since many GPs in Germany provide psychosomatic or psychotherapeutic care, this exclusion criterion would have substantially lowered the number of eligible GP practices. These GPs were thus not excluded from participation but their patients were only recruited following routine GP consultation, never following psychotherapeutic consultations.

#### Patients

Data on the primary and on most secondary outcomes was collected in tobacco smoking patients consecutively consulting their GP on data collection days by means of questionnaire-guided, face-to-face interviews immediately following GP consultation in the practice setting. Data collection was carried out by four trained part-time researchers. Per study cycle, each of them collected data in two or three practices. Prior to the consultation with the GP, patients were informed about, and invited to participate in the study by the researcher. At this time, all patients (independently of their smoking status) were invited to participate, and patients did not receive full information about the real purpose of the study (detailed information on blinding can be found in the corresponding paragraph). Following the consultation, the interviews were conducted in a separate room. All consenting patients were asked to answer questions on their sociodemographic characteristics and their current tobacco smoking status. Current tobacco smoking was defined as smoking cigarettes (including hand-rolled or self-stuffed) daily or occasionally, or any other combustible tobacco (pipe, cigars, cigarillos, shisha). For non-smokers, the interview ended at this point, whereas for current smokers the interview was continued with questions on their smoking behaviour, and on the outcomes of this study. In current smokers, the interview took approximately 10 to 15 minutes. The full baseline questionnaire can be found here: <u>osf.io/f2p7b/</u> (translated English version), <u>osf.io/7pmr5/</u> (original German version).

Patients younger than 18 years, those suffering from moderate or severe cognitive impairment, those with language barriers or of too low literacy to understand the patient informed consent form, or those who did not see their GP in person (e.g., just picking up a prescription) were ineligible to participate. Moreover, patients who participated in the first period of data collection prior to the GP training were ineligible to be interviewed again following the training. Although not explicitly mentioned in the protocol,^27^ patients using only electronic cigarettes or heated tobacco products were also not eligible to participate.

### Randomisation and masking

Cluster randomisation was used because the training was delivered at the practice level. Two small-group trainings – one per training method – were offered per study cycle, resulting in 12 trainings in total (six in ABC, six in 5As). GPs from the same practice were assigned to the same training. If a GP had to cancel the training, she or he was re-assigned to the same training including post-training data collection at the following study cycle. In group practices, only patients from GPs participating in the study were included. The minimum number of GPs to run the training was set at three, which occurred once. The maximum number was set at ten GPs, allowing intensive practical training. One training was conducted with 12 GPs due to group practices randomised to the same training.

To yield a maximum participation rate, three to four potential training dates were offered per cycle. GPs had to register for at least two dates to participate. Depending on how many GPs were available at least at two of the proposed training dates, two different methods of randomisation were applied:^27^

- Eight or more GPs: computer-generated block randomisation with permuted blocks of sizes two or four, prepared by an independent statistician (WV), and concealed from the study team.
- Fewer than eight GPs: randomisation by virtue of the GPs temporal availability, meaning that the two dates with most registrations were selected and, in a random order between the study cycles, one was assigned to be an ABC and the other to be a 5As training.

GPs could not be fully blinded to their training allocation, but no detailed information on both training methods or on group allocation was given until the end of the pre-training data collection had ended.^27^ Patients were blinded to the nature and aim of the study until the end of the data collection. The study was masked as a study on “physician-patient communication on health behaviour” in the initial informed-consent form. Following data collection, patients received full information on the purpose of the study, a strategy that was approved by the ethics committee. However, it cannot be fully excluded that patients talked to each other regarding the purpose of the study. The risk for this bias can assumed to be equally distributed across study arms and data collection periods.

It was not feasible to blind the researchers who collected the data to the GPs’ group allocation, but they were not actively involved in the trainings, and were alternately assigned to the practices for data collection as well as depending on the travel distance between a practice and their private residence.

### Procedures (Interventions)

In 2016, we developed a standardised 3.5h-training for GPs in delivering brief stop-smoking advice during routine practice consultations according to two different methodological methods: ABC^21^ and 5As^1^. The theoretical foundation of this training was the “COM-B” behaviour change model.^28^ The training was designed to address at least two components (**c**apability and **m**otivation) of the COM-B model which, according to the model, influence **b**ehaviour. Since the ABC method seems to be less difficult and less time consuming to apply for GPs, we assumed that the ABC training might also influence the third component “**o**pportunity” of the COM-B model. We used the Behaviour Change Techniques (BCT) Taxonomy^29^ to describe the active components of our training which might have the potential to alter the GPs’ behaviour. BCTs are reported in the study protocol.^27^ Trainings were always led by a senior researcher together with an experienced GP peer-trainer who both rotated between ABC and 5As trainings. Overall, three researchers and four GPs served as trainers. Each training started with an introductory lecture of about 60 min according to national and international guidelines on smoking cessation. This lecture included the latest evidence on tobacco addiction, effective smoking cessation treatments, details about the method of delivering stop-smoking advice (ABC or 5As), and reflexive group discussions on GPs’ experience with barriers to and facilitators of the provision of stop-smoking advice. The lecture was followed by 90 min of simulated role-plays, including moderated peer feedback, with professional actors trained in patients’ specific behaviour. GPs received one-page handouts on the structure of the respective method. In addition, and regardless of the method used, all GPs received information on evidence-based smoking cessation treatments, and a copy template with local outpatient programs, quit smoking websites and hotlines patients can be referred to. These handouts were developed as a result of the process evaluation following the pilot study.^27^ Participation in training was incentivised with five Continuing Medical Education credits.

### Outcome measures

#### Primary outcome

We published an overview of the pre-specified outcomes together with the statistical analyses plan: <u>osf.io/36kpc/</u> (version 3-3). The primary outcome was defined as the number of patients prior to and following the training who report the receipt of brief stop-smoking advice during the consultation with their GP, irrespective of the training method, out of the total number of patients who stated to be current smokers at the time of the consultation. Delivery of brief stop-smoking advice was assessed by asking the patient whether the GP urged him or her to quit smoking during this consultation.

#### Secondary outcomes

Most secondary outcomes refer to the patients’ consultation with their GP, and were measured together with the primary outcome. The following secondary outcomes were defined as the number of smoking patients prior to and following the training who, irrespective of the training method, reported the receipt of GP-delivered prescription or recommendation of: individual or group behavioural counselling in own practice or elsewhere, NRT, varenicline or bupropion, any pharmacotherapy (NRT, varenicline or bupropion), or a combination therapy of behavioural counselling and pharmacotherapy. Moreover, we aimed to directly compare the effectiveness of the ABC and the 5As method (interaction with pre vs. post measurement) by means of the primary and secondary outcomes.

At baseline, data on sociodemographic characteristics were collected on all patients. Among current smokers, further data were collected on smoking behaviour: average number of cigarettes (or, e.g., pipes, cigars) smoked per day, week or month (for occasional smokers), motivation to

(German version of the Motivation to Stop Smoking Scale, MTSS)^22^, and on urges to smoke (German version of the Strength of Urges to Smoke Scale, SUTS)^30^. Patients reporting the receipt of brief stop-smoking advice were asked about their satisfaction with that conversation; operationalised by ratings on a 6-point Likert scale ranging from 1 = “very satisfied” to 6 = “very dissatisfied”).

Further secondary outcomes were measured by means of postal follow-up questionnaires at week 4, 12, and 26 following the GP consultation. These data included attempts to quit smoking, the use of evidence-based or non-evidence-based smoking cessation methods (e.g., acupuncture, hypnosis), and point prevalence abstinence rates, but are subject to future analyses.

Data on GPs’ characteristics and of their practices (including age, gender, smoking status, professional experience, specialisation, and rural vs. urban practice location) were collected following randomisation. Data on short-term training effects on GP-reported attitude (motivation) towards, opportunity, knowledge on, and practical skills (capability) in the provision of brief advice to quit tobacco consumption were collected in accordance with the “COM-B” behaviour change model^28^ by means of a brief questionnaire prior to and immediately following the training.

### Statistical analysis

The primary outcome for this study was the percentage of smokers reporting the delivery of brief stop-smoking advice by their GP during routine consultation from prior to following the training. The sample size calculation was informed by data from our previous study of the German population^7^ showing that about 18% of smokers in Germany currently receive brief advice on smoking cessation (with/without treatment recommendation) during a consultation with their GP. From the pilot study^27^ we assumed that it would be feasible to recruit 48 GP practices during a period of about two years. Training GPs in either ABC or 5As was assumed to have a clinically relevant effect if it increases these rates by at least 10% (corresponding to an odds ratio of 1.77) between pre- and post-training. A simulation study showed that 16 patients (respectively eight prior to and eight following the training) per practice were needed to evaluate the primary outcome with a statistical power of at least 80%, and a total of 42 patients (respectively 21 prior to and 21 following the training) per practice were needed to evaluate the interaction effect between the time (pre-post training) and the group variable ABC vs. 5As (for which we assumed post-training percentages of 33% and 23%, respectively), resulting in a total study sample size of 2,016 patients (respectively 1,008 prior to and 1,008 following the training). Further details on the sample size calculation are reported in the protocol.^27^

#### Analyses of primary and secondary outcomes

To fully mask the statistician and the researchers involved in the statistical analyses of outcomes, we wrote the code for the present analyses prior to the analyses and based on a blinded dataset; i.e., with the values of the primary (pre vs. post training measurement) and secondary outcome variables (ABC vs. 5As training method) in a randomly shuffled order. All analyses were conducted using R version 3.6.1.^31^ The analysis plan (latest version 3.3) and R code (latest version 3.8) has been published on the Open Science Framework: <u>osf.io/36kpc/</u>, <u>osf.io/zurfq/</u>.

Data were structured hierarchically in clusters (= practices), with patients within these clusters. Since differences in rates of delivery of smoking cessation advice were expected among practices, mixed-effects logistic regression models were used to analyse the dichotomous primary outcome (received advice: yes vs. no), with a fixed effect for time (dichotomous: pre-vs. post training) and random effects for the practices and the time effect. The same model was applied to the secondary outcomes. All models were adjusted for potential confounders measured at baseline including patients’ age, sex, level of education, time spent with urges to smoke, and strength of urges to smoke.

In order to analyse differences between the ABC and 5As training, the group variable and its interaction with time (pre-post training) were added to the models as fixed effects. In both models, the time effect and the interaction were analysed by means of Wald-type tests (level of significance .05).

All participating patients were included in an intention-to-treat analysis. Since missing data on primary and secondary outcomes were very rare, no imputation methods were applied; see also paragraph “adherence to the protocol”. In contrast, missing data of potential confounding variables were imputed by using a multiple imputation approach, with missing data imputed by chained equations using the “mice-package”^32^ in R with m=20 imputed datasets and 10 iterations for each dataset. Results across the imputation datasets were pooled using Rubin’s rules.^33^ To examine the sensitivity of the results, an additional complete case analysis was performed for the primary outcome as well as for the interaction effect between group (ABC, 5As) and time (pre-post measurement).

### Adherence to the protocol

Smaller changes to the published study protocol regarding the data collection and GPs exclusion criteria are reported in the corresponding paragraphs throughout this manuscript.

All planned statistical analyses are reported in the study protocol and in the analysis plan published prior to conduction of the analyses (latest version 3.3, <u>osf.io/36kpc/</u>). A few changes made to these planned analyses including explanations are also provided within this document. We did not adjust the analyses for “motivation to stop smoking” since motivation was assessed following the consultation with the GP and might thus have been influenced by the behaviour of the GP during the consultation. Furthermore, we did not perform a complete case analysis for the primary outcome, since missing data were very rare (only 4 cases). For adjusted analyses, missing data of potential confounders were imputed to reduce the potential for bias compared to a complete case analysis.

Two additional secondary outcomes were assessed: an aggregate variable of all forms of patient-reported receipt of GP recommendation/prescription for pharmacotherapy (NRT, varenicline or bupropion) and the receipt of a combination therapy (pharmacotherapy and behavioural counselling). This decision was made since usage of stop-smoking medication is very low in Germany,^34^ and because combination therapy is recommended in the national clinical guideline.^3^

We further ran explorative subgroup analyses for the primary outcome with patient data (sex, level of education, and number of cigarettes smoked per day: ≤10 vs. >10) and GP data (sex, number of years in clinical practice, practice type, smoking status: ever vs. never smoker), but results are only reported if the interaction effect (subgroup variable by exposure variable) was statistically significant at p<0.05. Details on these group comparisons can be also found in the analyses plan (<u>osf.io/36kpc/</u>).

### Patient and public involvement

The trial procedure and all aspects of the GP training, including the case vignettes for the role-plays, were developed in an interdisciplinary team of healthcare researchers, GPs, and actors trained in patient-physician communication. During the pilot study,^27^ the training and methods of data collection were tested and reviewed, and feedback was obtained by means of a process evaluation in participating GPs regarding barriers and facilitators to transfer the content of the intervention into their daily practice routine. Methods of data collection in patients and comprehensibility of the questions to assess the study outcomes were also reviewed during the pilot study.

## RESULTS

**Figure 1** shows the trial flow of participating GPs and smoking patients. In total, 5,761 invitation letters were sent to addresses of GPs from single and group practices. A total of 106 practices responded, with at least one GP per practice generally interested to participate in the study. For 26 practices, no initiation telephone call could be scheduled with the GP in person, due to time constraints on either the GPs’ or the study centres’ side. Following a telephone call with GPs from the remaining practices, one GP did not meet the inclusion criteria and 21 GPs refused to participate due to time constraints, or an inability to provide a separate room for patient interviews. Finally, 58 practices with 78 GPs were eligible, provided informed consent, and received a study ident number. Two practices with three GPs withdrew before the start of the randomisation, hence a total of 56 practices (75 GPs) were randomly assigned to either an ABC or 5As training.

**Figure 1.**
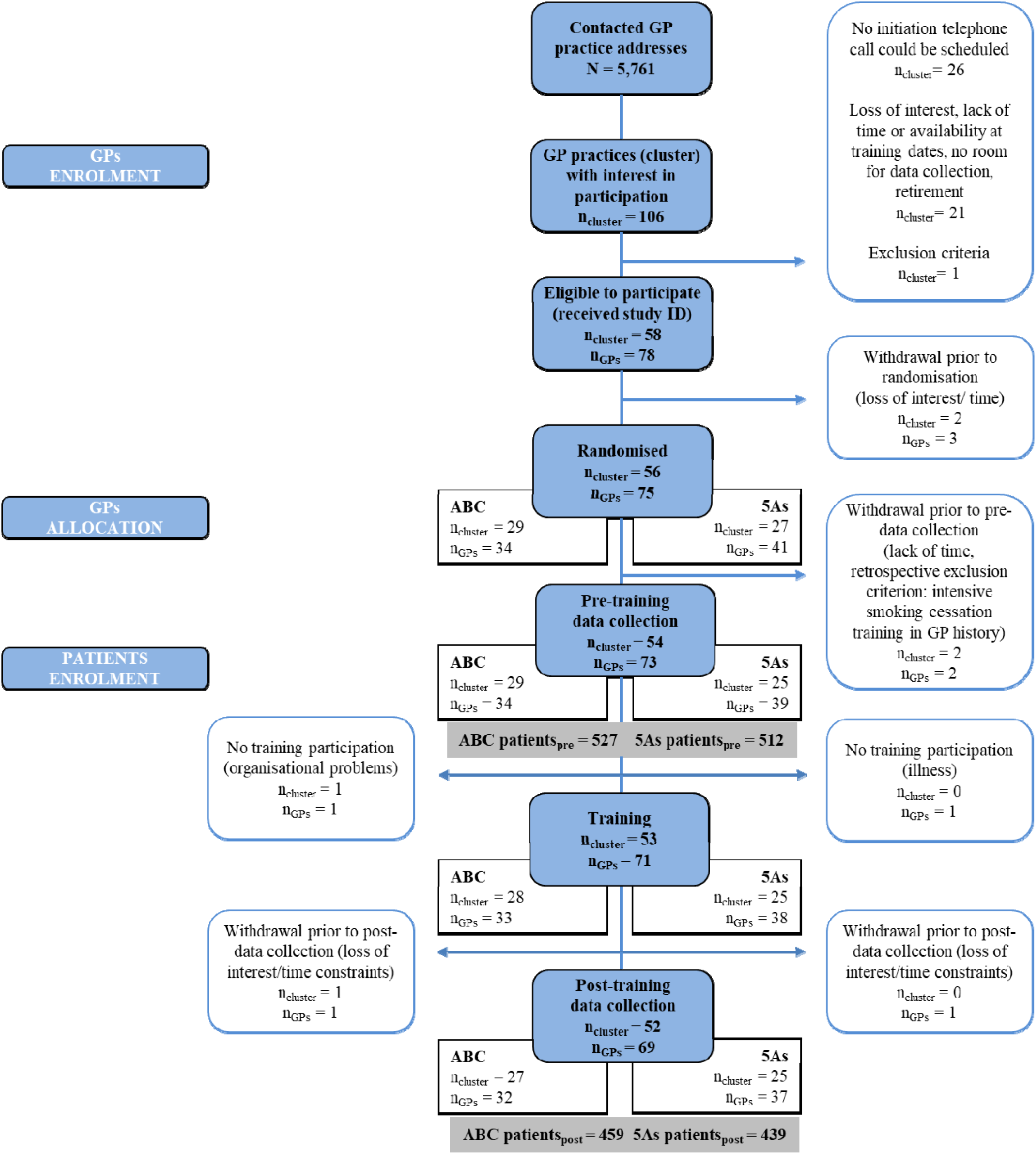
Consolidated Standards of Reporting Trials chart showing trial flow of general practices (cluster) with their general practitioners (GPs) and participating smoking patients by pre-training and post-training data collection period and study arm (ABC vs. 5As training); ID = identifier, n= number.

**Figure 2.**
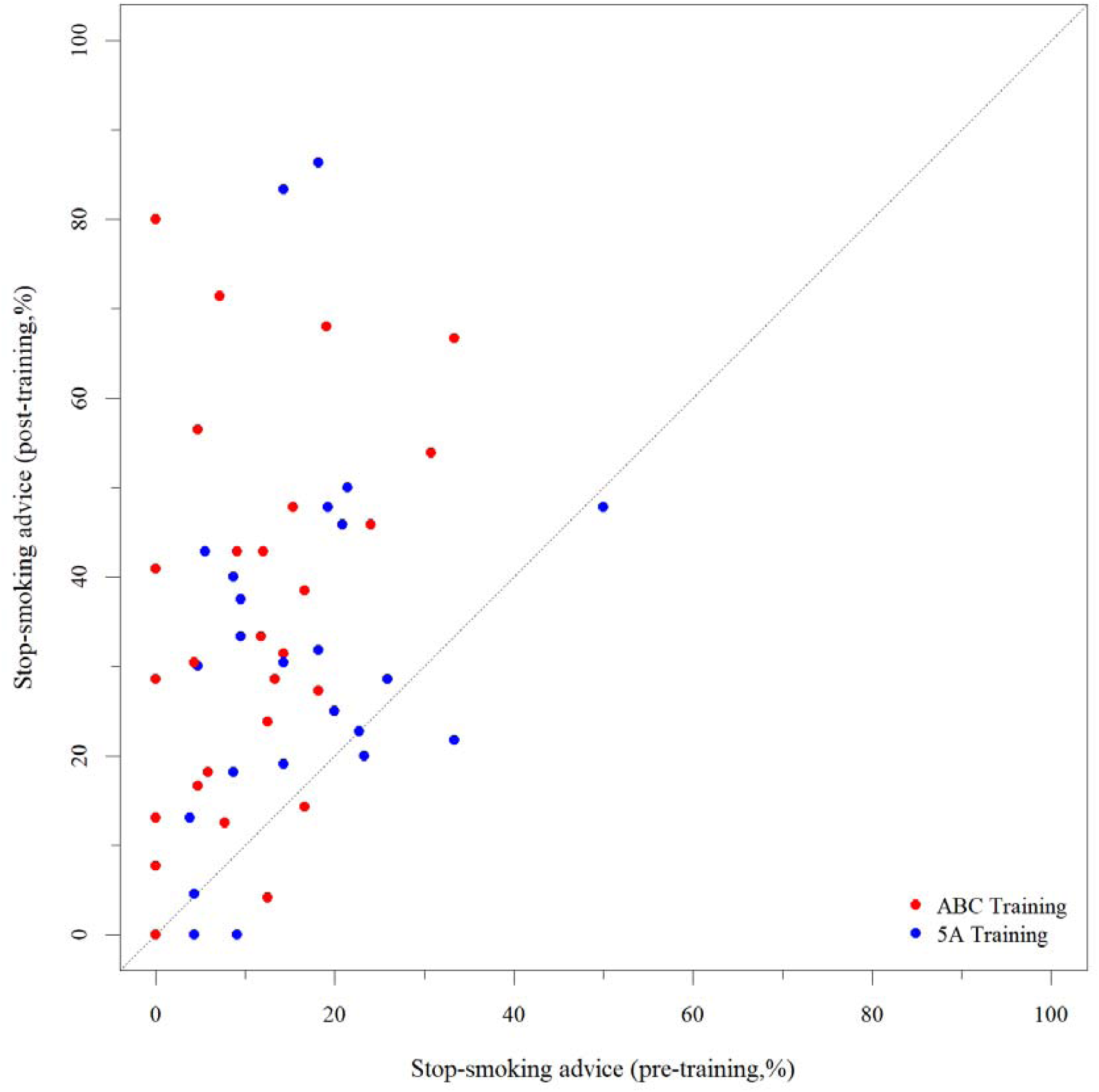
Scatter plot showing the relationship between the percentages of patients who reported the receipt of a stop-smoking advice delivered by their GP prior to the training (x-axis) and following the training (y-axis) by training group allocation of the GP.

During the process of the study, two practices per study arm had to be excluded because the GPs could not participate in the training or in the two periods of data collection due to acute illness, organisational difficulties, time constraints, and retrospective application of an exclusion criterion (**Figure 1**). A total of 52 practices (69 GPs) were finally included in the statistical analyses, a number which was slightly higher than originally intended. **Table 1** presents baseline sociodemographic and professional characteristics of this GP sample, stratified by training method.

**Table 1.** Baseline characteristics of general practitioners (GP), stratified by training method (N=69 GPs from 52 practices).

|  | **ABC**<br>**Training**<br>**(n = 32)** | **5As**<br>**Training**<br>**(n = 37)** | **Total**<br>**sample**<br>**(N = 69)** |
| --- | --- | --- | --- |
| Age, years (mean ± SD) | 51.5 ± 7.5 | 52.8 ± 8.0 | 52.2 ± 7.7 |
| Sex |  |  |  |
| Female | 56.3% (18) | 48.7% (18) | 52.2% (36) |
| Male | 43.8% (14) | 51.4% (19) | 47.8% (33) |
| Years since becoming physician (mean ± SD) | 22.6 ± 8.0 | 23.7 ± 9.0 | 26.1 ± 24.2 |
| Years since established in practice (mean ± SD) | 11.4 ± 8.6 | 13.5 ± 9.2 | 12.5 ± 8.9 |
| Type of GP practice |  |  |  |
| Single practice | 37.5% (12) | 21.6% (8) | 29.0% (20) |
| Any type of group practice | 62.5% (20) | 78.4% (29) | 31.0% (49) |
| Area where GP practice is located |  |  |  |
| Rural area (<20,000 inhabitants) | 3.1% (1) | 5.4% (2) | 4.4% (3) |
| Small city (>20,000 inhabitants) | 3.1% (1) | 5.4% (2) | 4.4% (3) |
| City (<100,000 inhabitants) | 31.3% (10) | 13.5% (5) | 21.7% (15) |
| Large city (>100,000 inhabitants) | 62.5% (20) | 75.7% (28) | 69.6% (48) |
| Ever participated in training on delivering smoking cessation before = Yes | 25.0% (8) | 21.6% (8) | 23.2% (16) |
| Current smoking status of GP |  |  |  |
| Daily smoker | 3.1% (1) | 0.0% (0) | 1.5% (1) |
| Occasional smoker | 15.6% (5) | 5.4% (2) | 10.1% (7) |
| Ex-smoker | 21.9% (7) | 32.4% (12) | 27.5% (19) |
| Never smoker | 59.4% (19) | 62.2% (23) | 60.9% (42) |

Data are presented as mean ± standard deviation (SD) or percentages (number, N); GP = general practitioner.

**Table 2** presents baseline sociodemographic of all patients of these GP practices who participated in the study and who were current tobacco smokers at that time, stratified by pre-post data collection period and by training method of the GP they had consulted. In total, 1,937 smoking patients participated of whom 1,037 patients were interviewed prior to the GP training and 898 patients following the GP training. The latter sample size was slightly lower than intended because some patients, i.e., those with acute diseases, visited their GP multiple times over several weeks, minimising the selection of unique patients within the second period of data collection.

**Table 2.** Baseline characteristics of all tobacco smoking patients, stratified by pre-post data collection period and by training method of the general practitioner they had consulted (N = 1,937).

|  | **Pre-**<br>**training**<br>**(n = 1,039)** | **Post-**<br>**training**<br>**(n = 898)** | **ABC**<br>**Training**<br>**(n = 986)** | **5As**<br>**Training**<br>**(n = 951)** | **Total**<br>**sample**<br>**(N =1,937)** |
| --- | --- | --- | --- | --- | --- |
| Age, years (mean ± SD) | 46.1 ± 16.1 | 46.0 ± 15.7 | 46.2 ± 16.0 | 45.9 ± 15.8 | 46.1 ± 15.9 |
| Sex |  |  |  |  |  |
| Female | 52.4% (544) | 52.1% (468) | 56.0% (552) | 48.4% (460) | 52.3% (1,012) |
| Male | 47.5% (493) | 47.7% (428) | 44.0% (434) | 51.2% (487) | 47.6% (921) |
| Level of education<sup>†</sup> |  |  |  |  |  |
| High school equiv. | 21.6% (224) | 22.8% (205) | 22.4% (221) | 21.9% (208) | 22.2% (429) |
| Adv. techn. college equiv. | 14.9% (155) | 13.2% (118) | 16.2% (160) | 11.9% (113) | 14.1% (273) |
| Secondary school equiv. | 29.7% (309) | 28.1% (252) | 29.3% (289) | 28.6% (272) | 29.0% (561) |
| Junior high school equiv. | 30.5% (317) | 32.3% (290) | 29.1% (287) | 33.7% (320) | 31.3% (607) |
| No qualification | 3.2% (33) | 3.6% (32) | 2.7% (27) | 4.0% (38) | 3.4% (65) |
| Cigarettes/day (mean ± SD) | 14.0 ± 9.3 | 13.6 ± 9.4 | 13.2 ± 9.2 | 14.5 ± 9.4 | 13.8 ± 9.3 |
| Time spend with urges to<br>smoke<sup>§a</sup> (mean ± SD) | 2.9 ± 1.5 | 3.0 ± 1.5 | 3.0 ± 1.5 | 2.9 ± 1.6 | 2.9 ± 1.5 |
| Strength of Urge to Smoke<br>(VRS)<sup>§b</sup> (mean ± SD) | 2.0 ± 0.9 | 2.1 ± 1.0 | 2.0 ± 0.9 | 2.1 ± 0.9 | 2.0 ± 0.9 |
| Motivation To Stop Smoking<br>(MRS)<sup>¥</sup> (mean ± SD) | 3.3 ± 1.8 | 3.3 ± 1.9 | 3.4 ± 1.8 | 3.1 ± 1.8 | 3.3 ± 1.8 |
| Satisfaction with conversation<br>on smoking with GP (if so)<sup>#</sup> | 2.0 ± 0.9 | 2.0 ± 0.9 | 2.0 ± 0.9 | 2.0 ± 0.9 | 2.0 ± 0.9 |

Data are presented as mean ± standard deviation (SD) or percentages (number, N). <sup>†</sup>German equivalents to education levels listed in table from highest to lowest: high school equivalent (“Allgemeine Hochschulreife”), advanced technical college equivalent (“Fachhochschulreife”), secondary school equivalent (“Realschulabschluss”), junior high school equivalent (“Hauptschulabschluss”), or no qualification; <sup>§</sup>VRS (“Verlangen zum Rauchen Skala”) = German version of the Strength of Urges to Smoke Scale (SUTS)<sup>30</sup>; SUTS<sup>§b</sup> is only posed to respondents who did not answer SUTS<sup>§a</sup> with “not at all”; <sup>¥</sup>MRS (“Motivation zum Rauchstopp Skaka”) = German version of the Motivation To Stop Scale (MTSS)<sup>22</sup>, <sup>#</sup>asked only to smoking patients who had a conversation on smoking with their GP (n=542) independently whether the patient reported the receipt of one of the outcomes, satisfaction was operationalised by ratings on a 6-point Likert scale ranging from 1 = “very satisfied” to 6 = “very dissatisfied; differences when calculating the total percentage can be explained by missing data on the respective variables.

Patients reporting the receipt of brief stop-smoking advice indicated that they were satisfied with that conversation with their GP, and no relevant differences regarding this satisfaction and patient characteristics were observed between the patient samples prior to and following the GP training and between the samples of the two GP training methods ABC and 5As (**Table 2**).

### Primary outcome

**Table 3** presents the results of the primary outcome analyses. The rates of patient-reported receipt of brief advice to quit smoking delivered by their GP increased from 13.1% (n = 136) prior to the training to 33.1% (n = 297) following the training (adjusted odds ratio (aOR) = 3.25, 95% confidence interval (CI) = 2.34 to 4.51, p<0.001). This result remained stable when using complete case data (aOR = 3.28, 95%CI = 2.35 to 4.59), in which patients with missing data on potential confounding variables were excluded (age: n = 2 (0.1%), level of education: n = 2 (0.1%), time spend with urges to smoke: n = 118 (6.1%), strength of urges to smoke^30^: n = 122 (6.3%)).

**Table 3.**
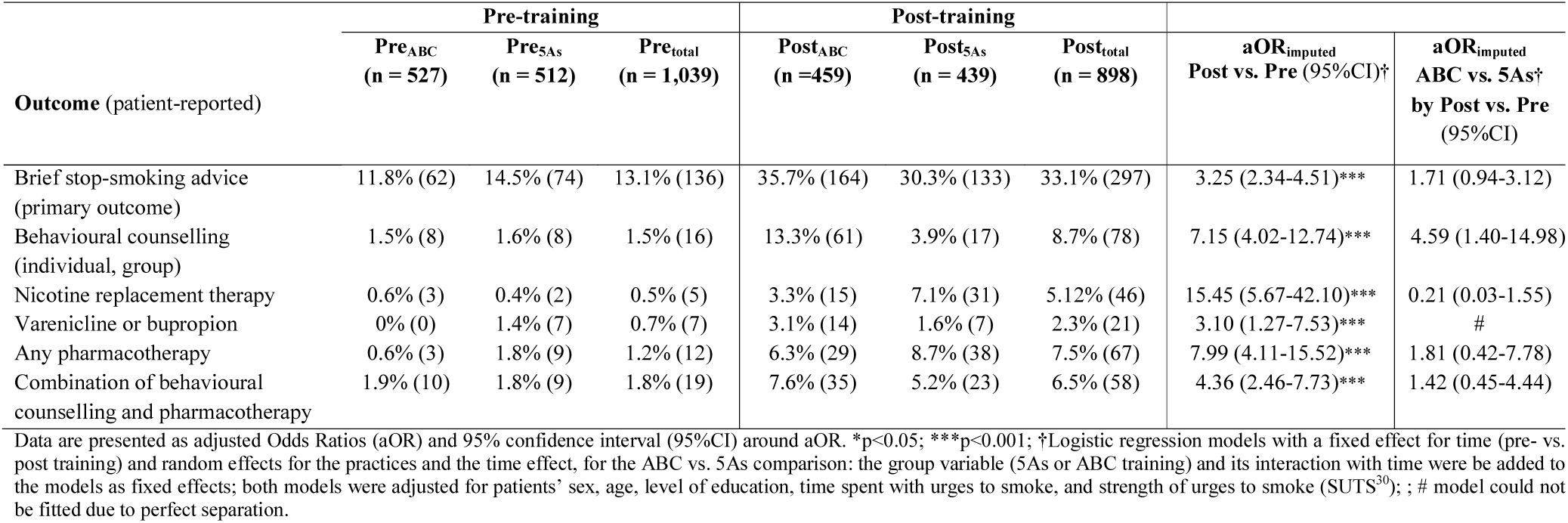
Patient-reported receipt of brief stop-smoking advice (primary outcome) and of recommendations/prescriptions of evidence-based treatment to quit smoking (secondary outcomes) delivered by their general practitioner (GP), stratified by pre-post data collection period and by training method of the GP they had consulted; and associations of these outcomes with training (pre vs. post) and with the interaction of training by training method (ABC/5As by pre/post) (N=1,937 smoking patients)

| Outcome (patient-reported) | Pre-training |  |  | Post-training |  |  | aOR<sub>imputed</sub><br>Post vs. Pre (95% CI)<sup>†</sup> | aOR<sub>imputed</sub><br>ABC vs. 5As<sup>†</sup><br>by Post vs. Pre<br>(95% CI) |
| --- | --- | --- | --- | --- | --- | --- | --- | --- |
|  | Pre<sub>ABC</sub><br>(n = 527) | Pre<sub>5As</sub><br>(n = 512) | Pre<sub>total</sub><br>(n = 1,039) | Post<sub>ABC</sub><br>(n = 459) | Post<sub>5As</sub><br>(n = 439) | Post<sub>total</sub><br>(n = 898) |  |  |
| Brief stop-smoking advice<br>(primary outcome) | 11.8% (62) | 14.5% (74) | 13.1% (136) | 35.7% (164) | 30.3% (133) | 33.1% (297) | 3.25 (2.34-4.51)*** | 1.71 (0.94-3.12) |
| Behavioural counselling<br>(individual, group) | 1.5% (8) | 1.6% (8) | 1.5% (16) | 13.3% (61) | 3.9% (17) | 8.7% (78) | 7.15 (4.02-12.74)*** | 4.59 (1.40-14.98) |
| Nicotine replacement therapy | 0.6% (3) | 0.4% (2) | 0.5% (5) | 3.3% (15) | 7.1% (31) | 5.12% (46) | 15.45 (5.67-42.10)*** | 0.21 (0.03-1.55) |
| Varenicline or bupropion | 0% (0) | 1.4% (7) | 0.7% (7) | 3.1% (14) | 1.6% (7) | 2.3% (21) | 3.10 (1.27-7.53)*** | # |
| Any pharmacotherapy | 0.6% (3) | 1.8% (9) | 1.2% (12) | 6.3% (29) | 8.7% (38) | 7.5% (67) | 7.99 (4.11-15.52)*** | 1.81 (0.42-7.78) |
| Combination of behavioural<br>counselling and pharmacotherapy | 1.9% (10) | 1.8% (9) | 1.8% (19) | 7.6% (35) | 5.2% (23) | 6.5% (58) | 4.36 (2.46-7.73)*** | 1.42 (0.45-4.44) |

Data are presented as adjusted Odds Ratios (aOR) and 95% confidence interval (95%CI) around aOR. \*p<0.05; \*\*\*p<0.001; <sup>†</sup>Logistic regression models with a fixed effect for time (pre- vs. post training) and random effects for the practices and the time effect, for the ABC vs. 5As comparison: the group variable (5As or ABC training) and its interaction with time were added to the models as fixed effects; both models were adjusted for patients' sex, age, level of education, time spent with urges to smoke, and strength of urges to smoke (SUTS<sup>30</sup>); ; # model could not be fitted due to perfect separation.

### Secondary outcomes

**Table 3** presents the results for the secondary outcomes. Overall, the patient-reported rates of GP-delivered recommendation/prescription of evidence-based treatment for smoking cessation were low prior to the training (<2%), but increased significantly for all types of evidence-based treatment after the training, including the combination of behavioural counselling and pharmacotherapy to quit smoking (aOR = 4.36, 95%CI = 2.46 to 7.73).

Regarding the comparison of the effectiveness of the two training methods (ABC vs. 5As) for the primary outcome, a higher increase in the rates of stop-smoking advice following the training was observed in patients whose GPs were trained according to ABC compared to those whose GPs were trained in 5As (aOR=1.71, 95%CI=0.94 to 3.12), although the difference failed to be statistically significant (p=0.08). This result remained stable when using complete case patient data for the analyses (OaR=1.69, 95%CI=0.91 to 3.12).

The increase in patient-reported rates of recommendations of, or referrals for, individual or group behavioural support delivered by the GPs following the training was higher in the ABC vs. 5As group (aOR=4.59, 95%CI=1.40 to 14.98). No such difference could be observed regarding the recommendation/prescription rates of stop-smoking medication or for the combination therapy (**Table 3**). One model (recommendation of varenicline or bupropion) could not be fitted due to perfect separation (prior to the training, no such recommendation was ever provided in the ABC group, increasing to 3.1% after the training, while the pre- and post-training percentages remained relatively stable at 1.4% and 1.6%, respectively, in the 5As group).

### Subgroup analyses

Explorative subgroup analyses for the primary outcome revealed a significant (p=0.025) interaction for the GPs’ number of years working in clinical practice (below group median number of years (≤ 12 years) vs. above group median number of years (>12 years)), showing a higher increase (from 11.3% to 36.1%) of GP-delivered stop-smoking advice following the GP training in patients who had visited a GP with less than 12 years of working experience (OR=4.63, 95%CI=2.93 to 7.33) compared to those who had visited a GP with more than 12 years in clinical practice (from 15.0% to 30.3%; OR=2.33, 95%CI=1.49 to 3.64). The mean difference in percentages between both groups prior to the training was not statistically significant (z-score: 1.388; p=0.165).

### GP-reported training effects

Data on short-term effects of the training on GPs’ attitude (motivation), knowledge and practical skills (capability), and opportunity to deliver brief stop-smoking are reported in **Supplementary Table S1**. Self-reported capability and opportunity in the provision of stop-smoking advice improved following the training, with all items increased by effect sizes between 0.58 and 2.84. The largest difference from prior to following the training was observed for self-reported knowledge of the steps of a structured brief stop-smoking advice (mean difference=2.84, p<.001, Cohen’s d=2.84). No such training effect could be observed for motivation, which was already high prior to the training (mean difference=0.07 and 0.10). Following the training, 91% (n=63) of the GPs agreed to implement brief stop-smoking advice more frequently in their daily practice (**Supplementary Table S2**). The majority (78%, n=55) estimated their learning growth following the training to be high or very high.

## DISCUSSION

### Principal findings of the study

In this cluster randomised controlled trial with pre-post data collection in 1,937 smoking patients, GPs’ participation in an 3.5h-training on the provision of brief stop-smoking advice according to the ABC or 5As method, including role-plays with professional actors and moderated by a senior researcher together with an experienced GP peer-trainer, was highly effective in increasing the rates of patient-reported receipt of such advice and recommendations of evidence-based smoking cessation treatment (behavioural, pharmacological, or combination of both) delivered by GPs.

Regarding the difference in the effectiveness of ABC and 5As, the increase of delivery of brief stop-smoking advice from prior to following the training was non-significantly higher in GPs trained in the ABC method compared to those trained in 5As. A significantly higher increase of patient-reported recommendation rates for behavioural support was observed in the ABC group. No such advantage was found for pharmacotherapy. However, the overall recommendation rates of such treatment were very low, resulting in large confidence intervals and inconclusive results.

### Strengths and limitations of study

Particular strengths of this study were the pragmatic “real-life” setting, the face-to-face data collection with low rates of missing data, and that data were assessed by means of patient reports, which is important since memory for medical information represents a necessary prerequisite for good adherence to recommended treatment.^35^ A detailed study protocol including results and lessons-learned from the pilot study has been published, and all statistical analyses were planned *a priori*. The analysis code was written on a dataset blinded for study arms and pre-post allocation, and published prior to the final analyses to yield maximum transparency and validity. The number of GP practices included was slightly higher than originally intended, resulting in adequately powered statistical analyses for the primary outcome. The recruitment rate of patients was marginally lower than expected since some patients recurrently visited their GP during both periods of data collection but could only participate once. For the ABC vs. 5As comparisons the analyses thus seem to be slightly underpowered.

Trial practices cannot be regarded as representative for all GP practices across Germany. However, North Rhine-Westphalia is the most populous German state, with a broad socioeconomic variability, and practices were located in urban and rural areas. A recent representative German population survey found rates (∼14%)^7^ of GP-delivered stop-smoking advice, strongly corresponding with the trial rates (∼13%) prior to the training, suggesting sound representativeness of the present data.

Patient-reported delivery of stop-smoking advice was chosen as primary outcome instead of validated abstinence, since abstinence strongly depends on the choice and use of smoking cessation treatment and can thus not reflect the direct effectiveness of the training. It can be seen as a major limitation of the present study that only short-term intervention effects were analysed. Whether or not long-term effects on GPs’ advice behaviour occur, and whether patients may benefit in terms of quit rates and success, thus needs to be determined. Only a small minority of GPs invited to participate in this study were enrolled. Hence, participating GPs may represent a selected group, suggested by high GP-reported interest in smoking cessation counselling prior to the training. However, pre-training data indicate that this high level of motivation is not related to an exceptional high activity level of delivering stop-smoking advice or treatment.

Data on main outcomes were patient-reported and may thus not reflect the actual performance of GPs. Moreover, the effectiveness of ABC vs. 5As can only be compared indirectly. In this pragmatic setting it could not be verified that GPs had effectively implemented the corresponding method. The most optimal method of data collection would have been audio or video observation of the consultation which was not feasible in this study. Data collection by means of face-to-face interviews following the consultation was the best alternative, also minimising recall bias. The presence of a study member itself might have influenced the GPs’ counselling performance. However, as described above, the rates of advice we found strongly match the rates of a recent population-based study,^7^ and substantial changes between the pre-to post-training data collection were observed despite the presence of a study team member in both periods.

As described in the study protocol^27^ and due to the pragmatic nature of this study, it was not possible to fully blind the researchers who conducted data collection. Several strategies were applied to reduce contamination. Researchers were required to avoid comments on the training and method while talking to GPs or patients, they were not actively involved in the trainings, and were assigned randomly or in relation to the distance of their private residence to the practices for data collection.

Although otherwise intended, in some practices the GPs directly referred smoking patients to the “study room”, which lead to better participation rates of patients than recruitment by the study member, but the influence on measured outcomes cannot be estimated. However, GPs acting accordingly kept this behaviour between pre- and post-training data collection.

### Comparison with other studies

Until to date, only few studies assessed the effectiveness of training GPs on the rates of delivery of smoking cessation advice or counselling.^15 17 18^ Verbiest et al.^18^ found that a one-hour 5As group training aimed at decreasing barriers to deliver brief advice to quit, increased the patient-reported frequency in which GPs asked about the smoking status, and the GP-reported frequency in which they advised smoking patients to quit. No effect was observed regarding the provision of evidence-based pharmacotherapy or regarding the arrangement of a follow-up contact (GP- and patient-reported).

Our training was developed based on the COM-B theory on behaviour change. Only the study of Girvalaki et al.^17^ also used a theory (Theory of Planned Behaviour) to guide the intervention design. Their pilot study found a full-day group training with two 3-hour refresher trainings to be highly effective in increasing the patient- and GP-reported rates of delivered smoking cessation counselling according to the 5As, including the discussion and prescription of stop-smoking medications. However, analyses were not adjusted for relevant patient characteristics and training duration was four times longer compared to our training, which may lower the GPs motivation to participate.

The study of Unrod et al., in contrast, showed that a 40 min individual 5As training session might be even effective in improving physicians’ implementation of all steps of the 5As in primary care.^16^ This study, as well as the study of McRobbie et al., which implemented a training of comparable length, did not particularly analyse prescription rates of smoking cessation treatment but discussions with the patients on cessation medication^16^ and referral rates to cessation services.^15 16^ However, in the German healthcare context, clinics providing behavioural group therapy are rare, and specialists’ stop smoking services such as in the United Kingdom^36^ do not exist. The GP therefore plays a central role in delivering stop-smoking advice and initiating effective treatment, which is why our training aimed at increasing the prescription and recommendation rates for evidence-based therapies.

The present study is the first comparing the effectiveness of the 5As vs. the much briefer ABC method on the performance of GPs of delivering brief stop-smoking advice. Higher delivery rates of behavioural support were observed in GPs trained according to ABC than according to 5As. Since the training content on smoking cessation treatment was standardised for both trainings, the most obvious reason for this difference might be that – according to our hypothesis – more smokers receive the “full dose” of ABC including “cessation support”. We can only hypothesise why this effect could not be observed for pharmacotherapy. In Germany, costs for behavioural support are at least partly reimbursed (50-75% of the costs) by health insurances, whereas pharmacotherapy is not reimbursed at all. This could affect the tendency to prescribe behavioural over pharmacotherapy. GPs also worry about the side-effects of stop-smoking medication, which might impact recommendation rates.^7^ Our training did not specifically address these reservations.

Comparable to a study of Bobak et al.^19^, who conducted a 3.5h-training for GP trainees, our training substantially increased the GP-reported capability and perceived opportunity to deliver stop-smoking advice during routine consultations. Although in the present study we measured only short-term effects, in the study of Bobak et al., this effect remained stable even at three-month follow-up.^19^

The present study revealed that GPs with fewer years of working experience in clinical practice seem to benefit more from the training intervention than those with more years in clinical practice. Reasons for this can only be hypothesised. It may be, for example, that GPs with less years of working in clinical practice are more open and have fewer difficulties to experiment with new, more patient-centred communication techniques, due to less well-established (and thus stiff) routines.

## Conclusions and policy implications

Our theory-based, 3.5h-training offers a highly effective strategy to improve the delivery of evidence-based smoking cessation advice in general practice. The data indicate that the ABC method might be more feasible to apply for GPs and thus might have a greater impact on the overall delivery of brief stop-smoking advice and behavioural smoking cessation treatment, and therefore may have the potential to decrease the national smoking prevalence. Efforts should be made to educate physicians on these simple counselling methods early on during professional training, and policy makers should be encouraged to implement the FCTC recommendations regarding the provision of counselling opportunities for smokers.

## Data Availability

The data underlying this study are third-party data (de-identified participant data, syntax of statistical analyses) and are available to researchers on reasonable request from the corresponding author. All proposals requesting data access will need to specify how it is planned to use the data, and all proposals will need approval of the trial co-investigator team before data release.

## Abbreviations

5A: Ask, advice, assess, assist, arrange
ABC: Ask, brief advice, cessation support
BCT: Behaviour Change Techniques
CI: Confidence Interval
COM-B: Capability-Opportunity-Motivation-Behaviour
FCTC: Framework Convention on Tobacco Control
GPs: General practitioner
HHU: Heinrich-Heine University
MTSS: Motivation to Stop Scale
NRT: Nicotine replacement therapy
SD: Standard deviation
SUTS: Strength of Urges to Smoke Scale

## Acknowledgements

The authors would like to thank all trial patients and general practitioners. In particular, we thank our GP peer trainers at the Institute of General Practice of the HHU Duesseldorf who were intensively involved in the development and provision of the training: Olaf Reddemann, Inken Blank, Detlef Maurer, and Elisabeth Gummersbach. We also thank our student assistants who contributed to the data entry and follow-up data collection: Esther Scholz, Sarah Fullenkamp, and Laura Schrobildgen. Stefanie Otten, trainer of the standardised patients, and her team (Brigitte Keldenich-Bergstein, Michael Hoch, and Georg Hoeren) is thanked for their professional and motivated assistance with the role-plays in preparation of and during the trainings.

## DECLARATIONS

### Contributors

DK conceived the study and acquired funding for the current study together with SK and VL, and co-wrote this manuscript. SK, VL, and DK developed and conducted the GP trainings. SK coordinated all study processes, and wrote the first draft of the current manuscript. WV, statistician, conducted a simulation study for the sample size calculation of the study and prepared the randomisation sequence. He also prepared the statistical analysis code, and advised on statistical analysis plans together with DK and SK, who conducted all analyses and interpreted the data. JH, DL, SKH, and CF were mainly involved in all study processes including recruitment, data collection, data entry and cleaning. OR, a general practitioner and peer trainer, was mainly involved in the final evaluation of the training manual and the didactic methods. SW, RW, TR, and HM gave valuable feedback at the time of designing the trial, and commented on and added to the present manuscript. All named authors contributed substantially to the manuscript and agreed on its final version. SK and DK are the guarantors. The corresponding author attests that all listed authors meet authorship criteria and that no others meeting the criteria have been omitted.

### Ethical approval

The study protocol was approved by the medical ethics committee at the Heinrich-Heine-University Düsseldorf, Germany (5999R).

### Transparency statement

The corresponding author (Sabrina Kastaun) affirms that the manuscript is an honest, accurate, and transparent account of the present study; that no important aspects of the study have been omitted; and that any discrepancies from the study as originally planned and registered have been explained.

### Data sharing

The data underlying this study are third-party data (de-identified participant data, syntax of statistical analyses) and are available to researchers on reasonable request from the corresponding author. The study protocol has been published.^27^ All proposals requesting data access will need to specify how it is planned to use the data, and all proposals will need approval of the trial co-investigator team before data release.

### Copyright/license for publication

The Corresponding Author has the right to grant on behalf of all authors and does grant on behalf of all authors, a worldwide licence to the Publishers and its licensees in perpetuity, in all forms, formats and media (whether known now or created in the future), to i) publish, reproduce, distribute, display and store the Contribution, ii) translate the Contribution into other languages, create adaptations, reprints, include within collections and create summaries, extracts and/or, abstracts of the Contribution, iii) create any other derivative work(s) based on the Contribution, iv) to exploit all subsidiary rights in the Contribution, v) the inclusion of electronic links from the Contribution to third party material where-ever it may be located; and, vi) licence any third party to do any or all of the above.

### Dissemination to participants and related patient and public communities

The study protocol is publicly available: https://bmcfampract.biomedcentral.com/articles/10.1186/s12875-019-0986-8. A preprint version of the study is publicly available on medRxiv: https://doi.org/10.1101/2020.03.26.20041491.

## Supplemental files

**Supplementary Table 1**

Changes in capability, opportunity and motivation to deliver brief stop-smoking advice to smoking patients from prior to following the training reported by 69 general practitioners (GPs) of 52 GP practices.

**Supplementary Table 2**

Relevance of the training content and learning curve reported by general practitioners (GPs) directly following the training (N=69 GPs from 52 GP practices).

**Supplementary Material 1:**

Original German version of the Baseline questionnaire for patients (BaselineSurvey_ABCII_German.pdf) can be downloaded here: <u>osf.io/7pmr5/</u>.

**Supplementary Material 2:**

Translated version of the Baseline questionnaire for patients into English (BaselineSurvey_ABCII_Engl.pdf) can be downloaded here: <u>osf.io/f2p7b/</u>.

## REFERENCES

1. Fiore MC, Jaen CR, Baker TB, et al. A Clinical Practice Guideline for Treating Tobacco Use and Dependence: 2008 Update - A US Public Health Service report. American Journal of Preventive Medicine 2008;35:158–76.

2. National Institute for Clinical Excellence (NICE). Smoking: acute, maternity and mental health services, Guidance PH48. 2013, https://www.nice.org.uk/guidance/ph48 (accessed 17 Mar 2020).

3. Arbeitsgemeinschaft der Wissenschaftlichen Medizinischen Fachgesellschaften (AWMF) S3 Guideline “Screening, Diagnostics, and Treatment of Harmful and Addictive Tobacco Use” [S3-Leitlinie “Screening, Diagnostik und Behandlung des schädlichen und abhängigen Tabakkonsums”]. AWMF-Register Nr. 076-006. 2015, http://www.awmf.org/leitlinien/detail/ll/076-006.html (accessed 07 Mar 2020).

4. Van Schayck OCP, Williams S, Barchilon V, et al. Treating tobacco dependence: guidance for primary care on life-saving interventions. Position statement of the IPCRG. npj Primary Care Respiratory Medicine 2017;27:38.

5. Stead LF, Koilpillai P, Fanshawe TR, et al. Combined pharmacotherapy and behavioural interventions for smoking cessation. Cochrane Database of Systematic Reviews 2016

6. Hartmann□Boyce J, Hong B, Livingstone□Banks J, et al. Additional behavioural support as an adjunct to pharmacotherapy for smoking cessation. Cochrane Database of Systematic Reviews 2019

7. Kastaun S, Kotz D. ärztliche Kurzberatung zur Tabakentwöhnung – Ergebnisse der DEBRA Studie. SUCHT 2019;65:34–41.

8. Strobel L, Schneider NK, Krampe H, et al. German medical students lack knowledge of how to treat smoking and problem drinking. Addiction 2012;107:1878–82.

9. Twardella D, Brenner H. Lack of training as a central barrier to the promotion of smoking cessation: a survey among general practitioners in Germany. European Journal of Public Health 2005;15:140–5.

10. Hoch E, Franke A, Sonntag H, et al. Raucherentwöhnung in der primärärztlichen Versorgung – Chance oder Fiktion? Ergebnisse der “Smoking and Nicotine Dependence Awareness and Screening (SNICAS)”-Studie. Suchtmedizin in Forschung und Praxis 2004;6:47–51.

11. Raupach T, Merker J, Hasenfuss G, et al. Knowledge gaps about smoking cessation in hospitalized patients and their doctors. European Journal of Cardiovascular Prevention and Rehabilitation 2011;18:334–41.

12. Cancer Research UK. Smoking Cessation in Primary Care: A cross-sectional survey of primary care health practitioners in the UK and the use of Very Brief Advice. 2019, https://www.cancerresearchuk.org/sites/default/files/tobacco_pc_report_to_publish_-_full12.pdf (accessed 02. December 2019).

13. Carson KV, Verbiest MEA, Crone MR, et al. Training health professionals in smoking cessation. Cochrane Database of Systematic Reviews 2012

14. Framework Convention on Tobacco Control (FCTC) Article 14 Guidelines. 2010, http://www.who.int/fctc/guidelines/adopted/article_14/en/ (accessed 28 Feb 2020).

15. McRobbie H, Hajek P, Feder G, et al. A cluster-randomised controlled trial of a brief training session to facilitate general practitioner referral to smoking cessation treatment. Tobacco Control 2008;17:173–6.

16. Unrod M, Smith M, Spring B, et al. Randomized Controlled Trial of a Computer-Based, Tailored Intervention to Increase Smoking Cessation Counseling by Primary Care Physicians. Journal of General Internal Medicine 2007;22:478–84.

17. Girvalaki C, Papadakis S, Vardavas C, et al. Training General Practitioners in Evidence-Based Tobacco Treatment: An Evaluation of the Tobacco Treatment Training Network in Crete (TiTAN-Crete) Intervention. Health Education & Behavior 2018:1090198118775481.

18. Verbiest ME, Crone MR, Scharloo M, et al. One-hour training for general practitioners in reducing the implementation gap of smoking cessation care: a cluster-randomized controlled trial.Nicotine & Tobacco Research 2014;16:1–10.

19. Bobak A, Raupach T. Effect of a short smoking cessation training session on smoking cessation behaviour and its determinants among GP trainees in England. Nicotine & Tobacco Research 2017

20. Twardella D, Brenner H. Effects of practitioner education, practitioner payment and reimbursement of patients’ drug costs on smoking cessation in primary care: a cluster randomised trial. Tobacco Control 2007;16:15–21.

21. Ministry of Health. The New Zealand Guidelines for Helping People to Stop Smoking. 2014, http://www.health.govt.nz/publication/new-zealand-guidelines-helping-people-stop-smoking (accessed 06 Mar 2020).

22. Kotz D, Brown J, West R. Predictive validity of the Motivation To Stop Scale (MTSS): a single-item measure of motivation to stop smoking. Drug and Alcohol Dependence 2013;128:15–9.

23. Hummel K, Brown J, Willemsen MC, et al. External validation of the Motivation To Stop Scale (MTSS): findings from the International Tobacco Control (ITC) Netherlands Survey. European Journal of Public Health 2017;27:129–34.

24. Martinez C, Castellano Y, Andres A, et al. Factors associated with implementation of the 5A’s smoking cessation model. Tobacco Induced Diseases 2017;15:41.

25. Bartsch AL, Harter M, Niedrich J, et al. A Systematic Literature Review of Self-Reported Smoking Cessation Counseling by Primary Care Physicians. PLoS One 2016;11:e0168482.

26. Park ER, Gareen IF, Japuntich S, et al. Primary care provider-delivered smoking cessation interventions and smoking cessation among participants in the national lung screening trial. JAMA Internal Medicine 2015;175:1509–16.

27. Kastaun S, Leve V, Hildebrandt J, et al. Effectiveness of training general practitioners to improve the implementation of brief stop-smoking advice in German primary care: study protocol of a pragmatic, 2-arm cluster randomised controlled trial (the ABCII trial). BMC Family Practice 2019;20:107.

28. Michie S, van Stralen MM, West R. The behaviour change wheel: a new method for characterising and designing behaviour change interventions. Implementation Science 2011;6:42.

29. Michie S, Richardson M, Johnston M, et al. The behavior change technique taxonomy (v1) of 93 hierarchically clustered techniques: building an international consensus for the reporting of behavior change interventions. Annals of Behavioral Medicine 2013;46:81–95.

30. Fidler JA, Shahab L, West R. Strength of urges to smoke as a measure of severity of cigarette dependence: comparison with the Fagerstrom Test for Nicotine Dependence and its components. Addiction 2011;106:631–8.

31. R Foundation for Statistical Computing. R: a language and environment for statistical computing.3.6.1 version. Vienna: Austria: R Core Team, 2013.

32. van Buuren S, Groothuis-Oudshoorn K. mice: Multivariate Imputation by Chained Equations in R.Journal of Statistical Software 2011;45:1–67.

33. Rubin DB. Multiple Imputation for Nonresponse in Surveys. New York: John Wiley and Sons 1987.

34. Kotz D, Batra A, Kastaun S. Smoking Cessation Attempts and Common Strategies Employed.Deutsches Arzteblatt International 2020;117:7–13.

35. Kessels RPC. Patients’ memory for medical information. Journal of the Royal Society of Medicine 2003;96:219–22.

36. Murray RL, McNeill A. Reducing the social gradient in smoking: initiatives in the United Kingdom.Drug and Alcohol Review 2012;31:693–7.

